# Antenatal surveillance of placental function using a wearable near infrared spectroscopy device with machine learning data interpretation

**DOI:** 10.64898/2026.03.12.26348170

**Authors:** Niccole Ranaei-Zamani, Zakaria Senousy, Temisan Ilukwe, Musa Talati, Sarah Johnson, Olivia Newth, Uzair Hakim, Darshana Gopal, Vatsla Dadhwal, Dimitrios Siassakos, Sara L Hillman, Hakim-Moulay Dehbi, Yevgeniya Kovalchuk, Anna L David, Ilias Tachtsidis, Subhabrata Mitra

**Affiliations:** Elizabeth Garrett Anderson Institute for Women’s Health, University College London, United Kingdom; Advanced Research Computing Centre, University College London, United Kingdom; Department of Medical Physics and Biomedical Engineering, University College London Hospital, United Kingdom; Department of Obstetrics, All India Institute of Medical Science, New Delhi, India; Department of Paediatric Pathology, Great Ormond Street Hospital for Children NHS Foundation Trust, London, United Kingdom; National Institute for Health and Care Research University College London Hospitals Biomedical Research Centre, London, United Kingdom; Comprehensive Clinical Trials Unit, University College London, United Kingdom

**Keywords:** Fetal monitoring, placental function, placental oxygenation, placental metabolism, near-infrared spectroscopy, fetal wellbeing, antenatal monitoring

## Abstract

**Background:** Placental dysfunction remains a leading cause of stillbirth and neonatal morbidity, yet current monitoring tools provide only indirect and intermittent measures of fetoplacental wellbeing. Near-infrared spectroscopy (NIRS) offers non-invasive, continuous monitoring of tissue oxygenation and metabolism.

**Objectives:** To develop a wearable NIRS system for placental monitoring (FetalSenseM v1 – FSM v1), investigate optical markers of placental oxygenation and metabolism in a population at high risk of adverse pregnancy outcomes such as stillbirth, and to apply machine learning analysis to develop a model for pregnancy outcome prediction.

**Study design:** In this prospective observational study, women with high-risk singleton pregnancies underwent antenatal placental NIRS monitoring for over 40 minutes. FSM v1 incorporates dual-source–detector separations and multiwavelength light sources to derive absolute placental oxygen saturation (PltO_2_) and relative cytochrome-c-oxidase (oxCCO) changes. FSM was placed on the abdominal wall following an ultrasound scan locating the placental position. Monte Carlo simulations were performed to estimate placental sensitivity, and a minimum placental sensitivity (MPS) threshold (>5%) defined a physiologically refined sub-cohort. Outcomes were classified using the In Utero near-miss criteria for stillbirth. Machine learning (ML) analysis evaluated 11 classifiers using nested stratified 5 × 4 cross-validation (5 outer folds for performance estimation and 4 inner folds for hyperparameter tuning).

**Results:** Seventy monitoring sessions from 58 participants were completed across gestational ages (25+2-41+1 weeks’ gestation); 33 recordings from 30 participants met MPS criteria. In the full cohort, mean PltO_2_ was 49.8% and was not related to gestational age or poor outcome based on near-miss stillbirth criteria. In the MPS sub-cohort, higher PltO_2_ was observed in severe fetal growth restriction (FGR) and lower PltO_2_ in gestational diabetes (both p=0.04). Hemodynamic-metabolic coupling (HbD-oxCCO semblance) was increased in severe FGR (p=0.0002). The best performing ML model (SVM) achieved a balanced accuracy of 78%, a recall (sensitivity) of 72% and a specificity of 84% under 5 x 4 nested cross-validation using the top 50 features. Feature importance analysis identified oxCCO-derived and haemodynamic–metabolic coupling features as dominant predictors, whereas static PltO_₂_ was non-discriminatory.

**Conclusion:** We describe the first wearable NIRS device to provide simultaneous non-invasive placental haemodynamic and metabolic monitoring. While static oxygenation indices lacked predictive value, ML analysis applied to dynamic NIRS features yielded accurate pregnancy outcome prediction, with metabolic signals emerging as key drivers. These findings support further development of wearable placental NIRS integrated with advanced analytics for antenatal surveillance.

**Condensation page:** *Tweetable statement:* A wearable placental near-infrared spectroscopy device enabled real-time monitoring of placental oxygenation and metabolism; machine learning of dynamic signals predicted risk of adverse pregnancy outcomes with 78% balanced accuracy.

**At a Glance:**

**A. Why was the study conducted?**
  - Current antenatal surveillance for assessment of fetal wellbeing is suboptimal.
  - We evaluated a wearable near-infrared spectroscopy device for real-time placental monitoring and outcome prediction
**B. What are the key findings?**
  - Machine learning applied to dynamic haemodynamic and metabolic optical signals of placental function identified pregnancies at risk with 78% balance accuracy
  - Placental oxygenation was higher in severe fetal growth restriction (FGR) and lower in participants with gestational diabetes (GDM)
**C. What does this study add to what is already known**
  - This is the first wearable near-infrared spectroscopy system to simultaneously monitor real-time changes in placental oxygenation and metabolism in vivo. This is also the first application of machine learning analysis to placental NIRS signals.
  - Dynamic features of placental metabolism and oxygenation levels may provide clinically meaningful placental biomarkers.

## Introduction

Despite major advances in antenatal care, the burden of adverse pregnancy outcomes such as stillbirth attributable to placental dysfunction remains high. Each year, more than two million babies are stillborn worldwide, while another 1.2 million babies born with hypoxic-ischaemic encephalopathy (HIE) ^1^. These complications are driven by impaired oxygen delivery and utilisation within the fetoplacental unit, yet the tools to assess fetal wellbeing currently available to clinicians largely provide surrogate or indirect measures.

Although current monitoring techniques are integral to antenatal care, their utility is restricted by technical and practical considerations. Ultrasound biometry and Doppler velocimetry can identify placental dysfunction, but their clinical effectiveness is resource-dependent, limited to hospital settings and prone to inter-operator error. Additionally, they only provide intermittent assessment of placental function and fetal wellbeing, leaving periods of vulnerability unmonitored. Cardiotocography (CTG) by contrast, is widely available but suffers from poor sensitivity and specificity for identifying fetal compromise, contributing to both missed hypoxia and high rates of unnecessary intervention ^2,3^. Maternal perception of fetal movement and blood-based biomarkers offer further adjuncts but remain limited to episodic assessments. Collectively, these approaches fail to provide continuous and sensitive placental monitoring, causing significant challenges for clinicians. Placental insufficiency is frequently undetected until too ate resulting in critical hypoxaemia and stillbirth, while equivocal findings may precipitate unnecessary obstetric intervention.

Near-infrared spectroscopy (NIRS) enables non-invasive, real-time assessment of tissue oxygenation and metabolism. This optical technique uses light in the near-infrared spectrum (650-950nm), a range in which biological tissues are relatively transparent. By emitting light at multiple wavelengths and detecting the portion that emerges after passing through tissue, NIRS exploits the distinct absorption spectra of chromophores to quantify their relative concentrations. In biological tissue, the principal absorbers of interest are oxygenated haemoglobin (HbO_2_), deoxygenated haemoglobin (HHb).When broadband or multi-wavelength systems are used, the changes in the redox state of the mitochondrial enzyme cytochrome-c-oxidase (oxCCO: the terminal electron acceptor in the mitochondrial respiratory chain), represent cellular oxygen utilisation. Therefore, NIRS can provide information on tissue oxygenation (StO_2_), blood volume (total haemoglobin, HbT), and oxygenation-related haemodynamic signals (haemoglobin difference, HbD) and metabolic activity (oxCCO).

Cerebral oximetry using continuous-wave (CW) NIRS is established in neonatal and adult critical care to monitor real-time changes in cerebral physiology and to guide interventions in preterm and critically ill infants^4,5^. This demonstrates both the safety and clinical value of NIRS to non-invasively continuously monitor tissues.

In obstetrics, however, NIRS is a research tool with early-stage trials in clinical settings. Recent evidence has demonstrated the technical feasibility of acquiring placental NIRS signals transabdominally both antenatally and during labour. Studies have been limited by small sample sizes, shallow penetration depths and restriction to anterior placentas^6–9,10^. Reported findings of placental NIRS studies are inconsistent, with both reduced and paradoxically elevated placental oxygenation described in pregnancies complicated by fetal growth restriction (FGR) or placental insufficiency^6–13^. This variability likely arises from differences in instrumentation, measurement depth, acquisition duration, signal processing and study design.

To overcome these challenges, we first optimised a time domain NIRS system developed by the UCL Biomedical Optics Research laboratory (MAESTROS II, an in-house built multiwavelength time-domain (TD) NIRS system)^14^, making a series of hardware modifications to enable simultaneous in-vivo measurement of placental haemodynamics and mitochondrial metabolism for the first time ^15^. We established normal ranges of expected values of placental oxygenation (PltO_2_) and metabolic function (as HbD:oxCCO semblance, a phase relationship between a haemodynamic oxygen-related signal and mitochondrial function) across gestational age in pregnancies with normal outcomes^16^.

The MAESTROS platform is a large and complex research device and has limitations for routine clinical use so to develop a small wearable device for placental function monitoring, we investigated the use of continuous-wave (CW) technology. Unlike TD-NIRS, conventional CW-NIRS is limited to tracking relative changes in chromophore concentrations. FetalSenseM version 1 (FSM v1) is a CW-NIRS, multiwavelength system incorporating dual source-detector separations (3 and 5cm). FSM v1 represents a wearable-format prototype rather than a fully wireless device^17^*l*^17^. UCLn algorithm is used to resolve relative concentration changes of the individual chromophores, while spatially resolved spectroscopy (SRS) and dual slope (DS) analysis provide absolute values of PltO_2_. By integrating these analytic approaches, FSM v1 is, to our knowledge, the first wearable prototype CW-NIRS device capable of delivering real-time information regarding changes in placental oxygenation (PltO_2_) with metabolic indices (oxCCO).

While NIRS provides rich physiological information, interpretation of these high-dimensional signals is complex. Conventional analyses typically rely on mean oxygenation values or simple group comparisons, which may not capture the dynamic interplay and nuances between haemodynamic and metabolic variables. Machine learning (ML) can extract predictive patterns from NIRS data, integrating multiple features simultaneously and learning non-linear relationships inaccessible to traditional approaches. Although ML has been applied to other obstetric monitoring modalities such as CTG, no prior studies have investigated ML analysis of placental NIRS^18^.

## Materials and methods

We conducted a single-centre, prospective observational study at University College London Hospital (UCLH) with clinical recruitment between May 2024 and March 2025. This study was approved by the East of England: Essex Research Ethics Committee (REC reference: 23/EE/0077; IRAS Project ID: 325344). All participants provided written informed consent prior to participation. The study was conducted in accordance with the Declaration of Helsinki.

### Participants

Eligibility criteria included participants over 18 years of age with singleton pregnancies with established risk factors for placental insufficiency, such as hypertensive disorders of pregnancy (essential hypertension, pregnancy-induced hypertension (PIH) or pre-eclampsia (PET)), diabetes (type 1, type 2 or gestational) and fetal growth restriction (FGR)/small-for-gestational age (SGA)^19^. Exclusion criteria were multiple pregnancy, pregnancies affected by known major fetal structural or genetic abnormalities.

### Monitoring protocol

Placental location was determined by ultrasound before optical monitoring. Abdominal layer (skin, adipose and composite muscle (rectus muscles and myometrium)) measurements were taken to determine placental depth. The skin tone of all participants was documented using the Fitzpatrick scale. Optical monitoring was performed for approximately 40 minutes, transabdominally after placing FSM over the placental site, with the probe secured using a CTG belt. The abdomen was covered using an opaque cloth during monitoring to shield FSM v1 from ambient light.

Clinical data from electronic healthcare records (EPIC). were entered into a secure REDCap database. Pregnancy outcomes were classified using the ‘Near-miss criteria for stillbirth’ outcome criteria developed by the In Utero Wellcome Leap consortium^21^. Adverse outcomes included stillbirth, high-grade maternal-vascular malperfusion (MVM) or fetal vascular malperfusion (FVM) on placental histopathology, late fetal loss (20-28 weeks’ gestation), severe pre-eclampsia (PET), neonatal cardiopulmonary resuscitation, arterial pH <7.1, 5-minute APGAR <7 and severe fetal growth restriction (FGR). Severe FGR was diagnosed if one of the following criteria was fulfilled: antenatally diagnosed absent or reversed end-diastolic flow in the umbilical artery, birth weight <10^th^ centile (INTERGROWTH) with abnormal antenatal Doppler or birth weight <3^rd^ centile (INTERGROWTH)^21^.

### Development of FetalSenseM v1 (FSM v1)

FSM v1 is a custom-built portable multiwavelength CW-NIRS device designed specifically for antenatal monitoring of placental oxygenation and metabolism. The device incorporates two light-emitting diode (LED) sources and two photodiode detectors, with source-detector separations of 3cm and 5cm to provide differential sensitivity to superficial and deeper tissues. Each source sequentially emits light at five discrete wavelengths (780, 810, 830, 850 and 890 nm), chosen to optimise sensitivity to haemoglobin and oxCCO absorption features^17^.

The optoelectronic components were integrated into a portable control unit with real-time data acquisition capability. The probe housing was ergonomically designed for placement on the maternal abdomen, with soft biocompatible materials to maximise comfort during prolonged monitoring. A custom data acquisition board managed LED driving, synchronous detection, and analogue-to-digital conversion. Raw intensity signals were digitised and streamed via USB cable to a secure laptop interface for storage and subsequent analysis.

### Phantom development

To validate the performance of FSM v1, a series of optical phantom studies were undertaken. Simple tissue-equivalent phantoms with defined absorption and scattering properties were constructed to mimic the optical characteristics of maternal abdominal tissues. These were used to test signal stability, noise levels, and the ability of the system to recover haemoglobin concentration changes under controlled conditions.

In addition, Monte Carlo simulations of photon transport were performed to evaluate the depth sensitivity of different abdominal layers using the dual source–detector configuration (3 cm and 5 cm separations), as described by *Caredda et al*^20^. These simulations confirmed that the 5 cm channel provided greater sensitivity to deeper structures, consistent with the expected location of the placenta, while the 3 cm channel was more sensitive to superficial layers. Together, these phantom experiments and simulations established the feasibility of using FSM v1 for transabdominal monitoring of placental oxygenation and metabolism.

### Optical algorithms

The UCLn algorithm was used to resolve relative concentration changes in individual chromophores (HbO_2_, HHb, and oxCCO), using a multi-wavelength modified Beer-Lambert Law approach enabling combined haemodynamic-metabolic analysis ^22^. PltO_2_ was estimated using spatially resolved spectroscopy (SRS) and dual slope (DS) analysis, which exploits differential light attenuation across source-detector separations to derive absolute tissue oxygen saturation. In contrast, Data quality was ensured by excluding recordings with an inadequate signal-to-noise ratio and by filtering segments affected by motion artefact.

### Wavelet analysis

Wavelet analysis was used to examine the dynamic relationship between haemodynamic oxygen-related changes and placental mitochondrial metabolism. Specifically, we analysed the coupling between HbD (a haemodynamic oxygen-related marker) and oxCCO (a marker of metabolic function) using wavelet semblance. Wavelet analysis enables time-frequency decomposition of non-stationary physiological signals, allowing assessment of whether fluctuations in. HbD and oxCCO occur in phase, out of phase or independently across different frequency bands. Continuous wavelet transforms were applied to HbD and oxCCO time series, and wavelet semblance was calculated to quantify their phase relationship, with values ranging from -1 (out of phase) to +1 (in phase). In neonatal hypoxic-ischaemic brain injury, a shift towards higher (more in-phase) haemodynamic-metabolic semblance has been interpreted as deranged or passive metabolic autoregulation and is associated with poor outcomes^5^. We applied the same framework here to interrogate placental haemodynamic-metabolic regulation in vivo.

### Signal processing

Raw voltage data from FSM v1 were converted into chromophore concentration changes and oxygenation indices using a semi-automated signal processing pipeline implemented via a cloud-linked interactive dashboard^22^. The dashboard enables standardized ingestion of raw voltage data, visualization of raw and processed signals, automated data quality assessment and user-supervised artefact cleaning. Signals were temporally resampled to a uniform sampling rate (1Hz), underwent automated quality control and filtering to minimise noise and motion artefact, and were then converted into chromophore concentration changes and PltO_2_ indices. This framework ensured consistent, transparent and reproducible signal processing across all recordings prior to downstream analysis.

### Monte Carlo simulation

A light propagation modelling of maternal abdomen was developed as a four-layer volume (skin, adipose tissue, muscle, and placenta)^20^. Measurements of skin tones and thickness of the skin, adipose tissue, and muscle layers were obtained from individual participants to calculate placental sensitivity. A minimum placental sensitivity (MPS) threshold was set at over 5% to identify a sub-cohort with definite placental contribution.

### Machine learning (ML) analysis

The FSM v1 dataset was analysed using a supervised machine learning pipeline implemented at the UCL Advanced Research Computing (ARC) Centre (Figure 2). Since raw time-series data frequently exhibit noise and missing values arising from device artefacts and external disturbances, pre-processing was performed to improve data quality before modelling. This included signal standardisation; temporal alignment of haemodynamic and metabolic channels; imputation of missing values using linear interpolation and forward and backward filling; and outlier detection and removal using Density-Based Spatial Clustering of Applications with Noise (DBSCAN).

**Figure 1.**
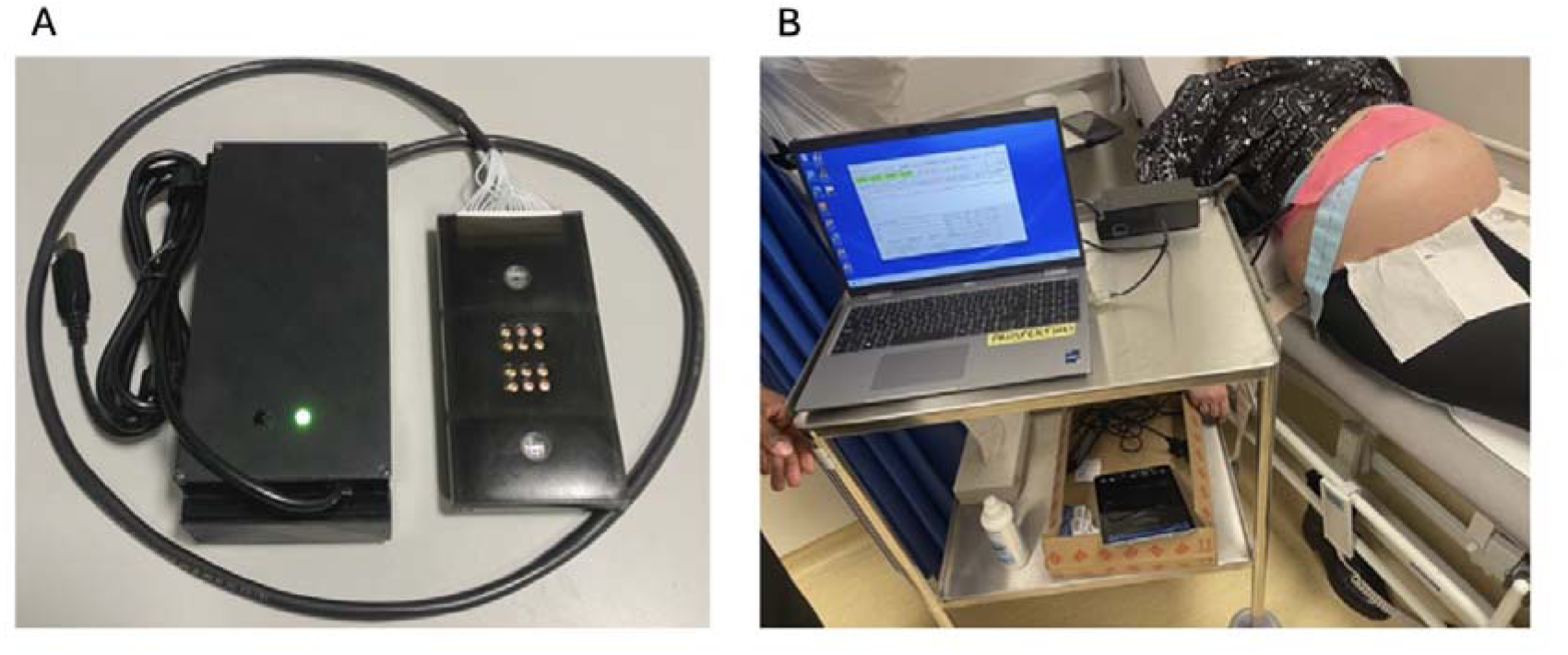
The first prototype of FetalSenseM (FSM v1) (A). FSM v1 is being used in the antenatal clinic for the study monitoring (B).

**Figure 2.**
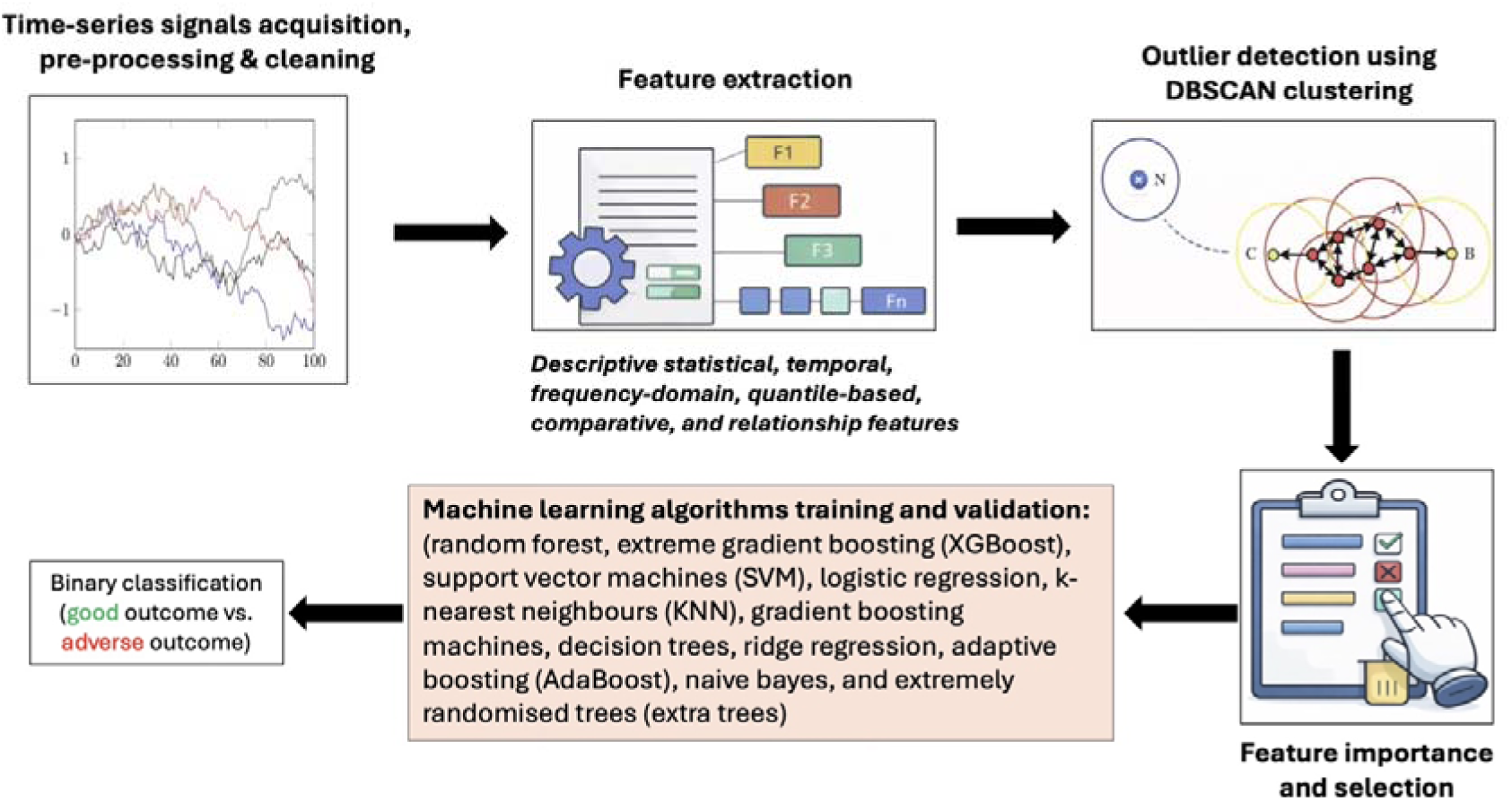
Flowchart indicating different steps of the machine learning pipeline

A total of 328 features serving as inputs into ML classifiers were engineered based on haemodynamic variables (HbO_2_, HHb, HbT, HbD and PltO_2_) and metabolic indices (oxCCO), together with frequency-domain and wavelet-derived measures to capture dynamic signal characteristics. Feature importance was evaluated using random forest^23^, and the top-ranking features were retained for subsequent modelling to reduce dimensionality and mitigate multicollinearity; classifiers were then applied across a series of top-*k* feature subsets (top 5, 10, 15, etc., up to 100 features), with the number of features yielding best performance for each classifier varying between 20 and 50.

A set of 11 classifiers was trained using the selected feature subsets, including random forest, extreme gradient boosting (XGBoost), support vector machine (SVM), logistic regression, k-nearest neighbours (KNN), gradient boosting machines, decision trees, ridge regression, adaptive boosting AdaBoost), naive bayes, and extremely randomized trees (extra trees). Model interpretability was investigated through feature-importance analysis, with variables ranked using random forest feature importance to quantify the contribution of individual haemodynamic and metabolic features.

Model training and evaluation was based on 5 x 4 nested cross-validation, whereby inner folds were used for model training and hyperparameter tuning, while outer test folds (i.e. data unseen by the trained model) were used to evaluate model’s performance, with this train-test process being repeated 5 times. The hyperparameter tuning was conducted using the Optuna framework^24^.

## Results

After assessing data quality and removing outliers based on the DBSCAN algorithm, NIRS datasets from 58 participants (70 sessions) were included for the analysis. A sub-cohort analysis was also performed with the Minimum Placental Threshold (MPS), set at over 5%. The sub-cohort consisted of datasets from 30 participants (33 sessions). Mean depth from skin-to-placenta was 23.61 mm (±8mm) for the full cohort and 16.48mm (±3.4mm) in the MPS sub-cohort.

### Participant characteristics

### Placental oxygenation and outcomes

The mean PltO_₂_ across all FSM datasets was 49.83% (±8.36%), and 51.24% (±5.37%) in the MPS sub-cohort. There was no significant change in PltO_₂_ with advancing gestation in either cohort (full cohort p=0.28, MPS sub-cohort p=0.72) (Figure 3).

**Figure 3.**
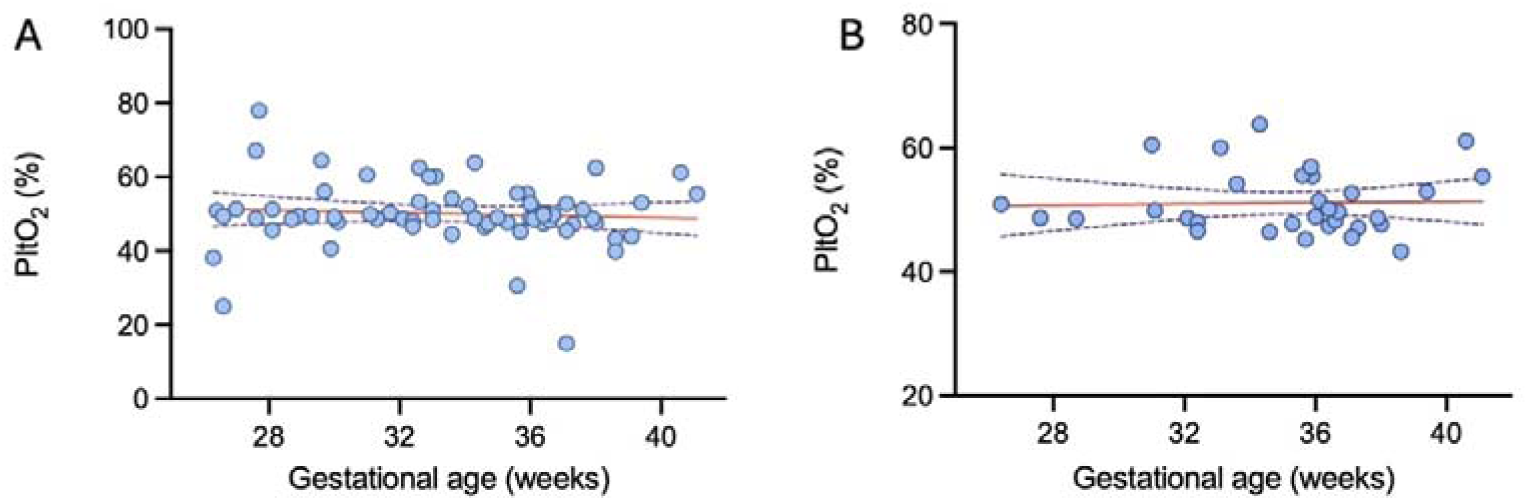
Linear regression analysis of mean PltO_2_ across gestational age in the full cohort (p=0.28) (A) and the MPS sub-cohort (p=0.72) (B).

In the full cohort, 28 (48.2%) pregnancies had a good outcome and 30 (51.7%) had an adverse outcome (positive for near-miss stillbirth criteria). There was no difference in mean PltO_2_ in pregnancies with good (49.73% ± 6.37%) or adverse outcomes (50.29% ± 10.56%) (p=0.78). There was no statistically significant difference in mean PltO_2_ in anterior, posterior or fundal placentas (p=0.31). Aetiologies associated with adverse outcomes are described in Table 1. A linear mixed-effects model with random intercept was used for comparison as some participants contributed to more than one study session. Pregnancy outcome was not associated with any differences in mean PltO_2_ (ß 0.15 for pregnancy outcome, 95% CI -4.86 -5.16, adjusted p=0.95). Gestational age at assessment was also not associated with mean PltO_2_ (ß -0.16 per gestational age week, 95% CI -0.75 – 0.42, p=0.58). The intraclass correlation coefficient (ICC) was negligible (0.002), and results were materially unchanged with models treating observations as independent. Therefore, subsequent analyses are presented using conventional regression models.

**Table 1.**
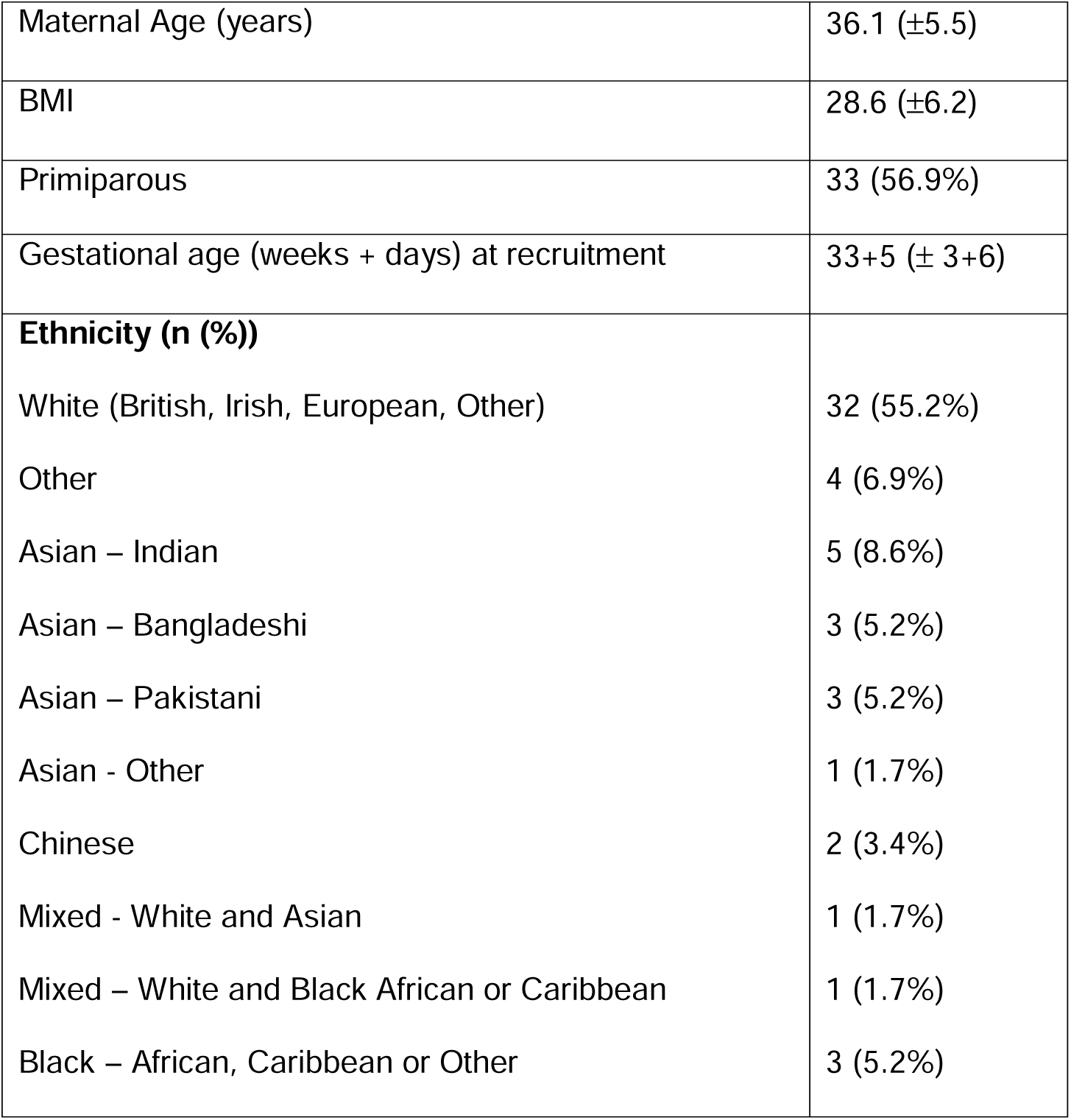
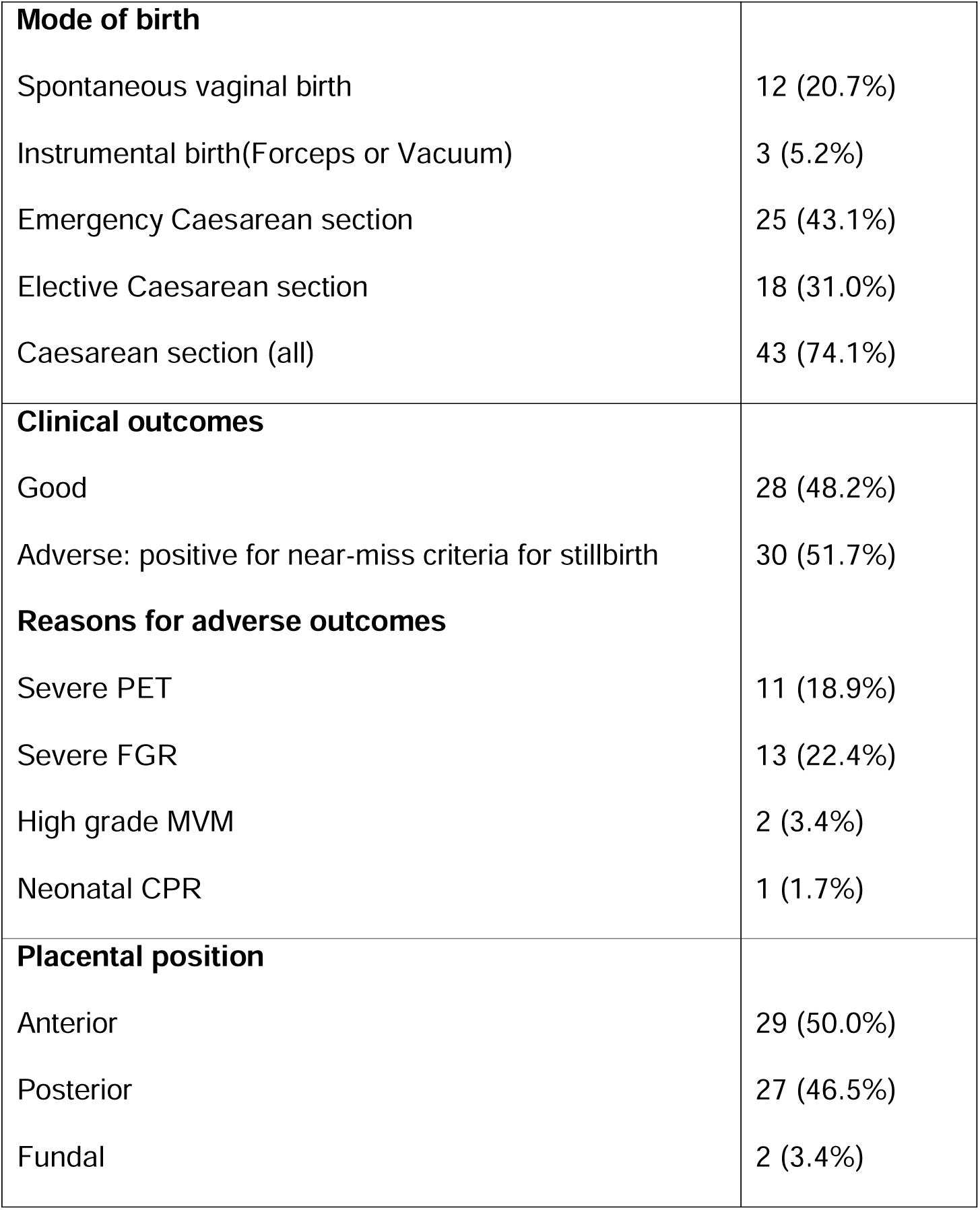
Study participant characteristics for the full participant cohort (n=58). PET: pre-eclampsia; FGR: fetal growth restriction; MVM: maternal villous malperfusion in the placenta; CPR: cardiopulmonary resuscitation.

In the MPS sub-cohort, 22 pregnancies had a good outcome, and 8 pregnancies had an adverse outcome. Although higher mean PltO_2_ values were noted in those with adverse outcomes (53.13% ± 6.04) compared with those with good outcomes (50.47±4.64), this did not reach statistical significance (p=0.28). In the MPS cohort, there was no statistically significant difference in mean PltO_2_ in anterior, posterior or fundal placentas (p=0.91). In the analysis of the full cohort, there were no statistically significant differences in mean PltO_2_ in participants with PET (49.30% ± 8.17%) compared with those without PET (50.36% ± 9.19) (p=0.64) or with diabetes (GDM, T1DM and T2DM) (49.48% ± 9.06%) compared with those without diabetes (50.36% ± 9.19%) (p=0.64). Higher mean PltO_2_ values were noted in participants with severe FGR (55.85% ± 6.14) compared with those without severe FGR (49.57% ± 8.89), although this did not quite reach statistical significance (p=0.08). In the MPS cohort analysis, there was no statistically significant difference in mean PltO_₂_ values in participants with PET (49.64% ± 2.78%) compared with those without PET (51.50% ± 5.48%) (p=0.23). However, those with GDM had a significantly lower mean PltO_2_ (49.61% ± 4.04%), compared with those without (53.73% ± 5.72%) (p=0.04) (Figure 4A). Participants with severe FGR had a significantly higher mean PltO_2_ (59.79% ± 4.04%), than participants without FGR (50.26% ± 4.36%) (p=0.04) (Figure 4B).

**Figure 4.**
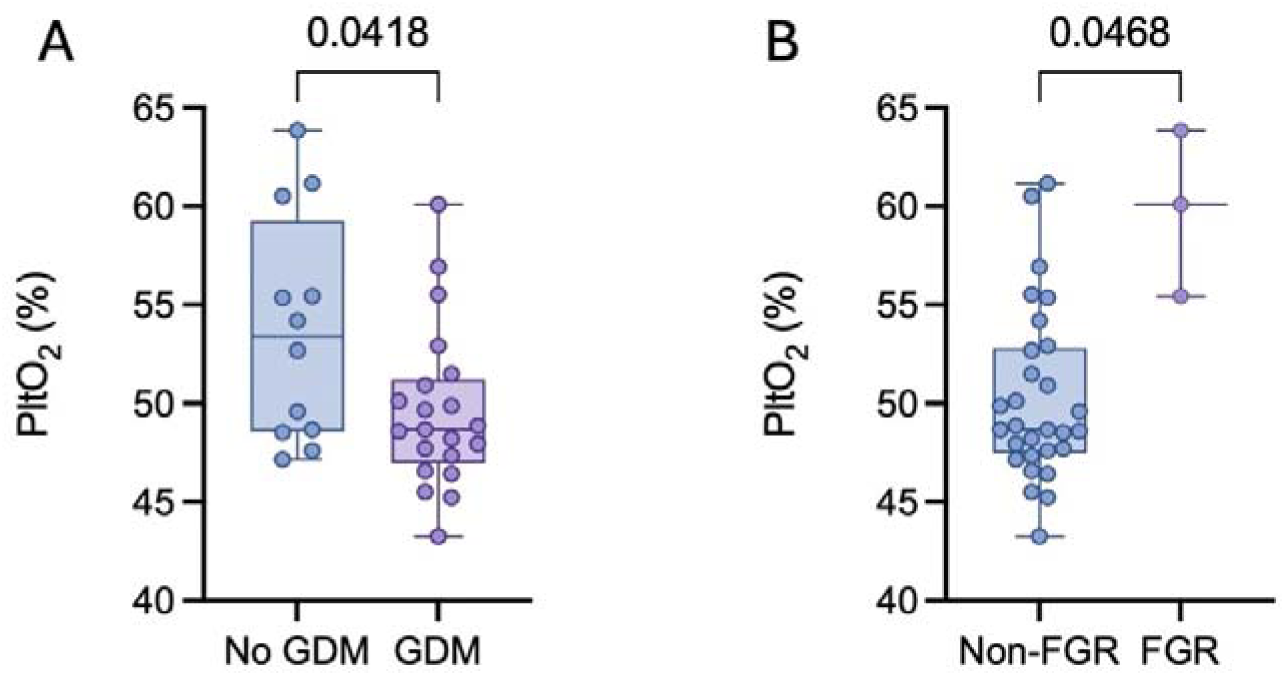
Mean PltO_2_ in pregnancies complicated by gestational diabetes and those without (p=0.0418) (A) and by severe-FGR and without (p=0.0468) (B). GDM: gestational diabetes; FGR: fetal growth restriction

### Wavelet analysis

Wavelet semblance analysis was performed to examine the relationship between haemodynamic and metabolic signals across frequency bands. There was no significant association between HbD:oxCCO or HbT:oxCCO semblance and gestational age in either the full frequency spectrum or slow-wave frequency range (all p>0.05). There were no differences in mean HbD:oxCCO values in participants with normal or adverse outcomes (0.06 ± 0.34 vs 0.05 ± 0.34, p=0.90). Higher HbD:oxCCO values were noted in severe FGR pregnancies (0.18 ± 0.27) however, this did not reach statistical significance when compared with values for normal outcomes (add in normal pregnancy results here p=0.68). There were no statistically significant differences in pregnancies complicated by PET (p=0.59) or GDM (p=0.99).

In the MPS sub-cohort, there was no significant association between HbD:oxCCO or HbT:oxCCO semblance and gestational age in either the full frequency spectrum or slow-wave frequency range (all p>0.05) (Figure 5a). In FGR pregnancies, HbD:oxCCO semblance values were higher (p=0.0002). There were no statistically significant differences in pregnancies complicated by PET (p=0.5571) or GDM (p=0.2642).

**Figure 5.**
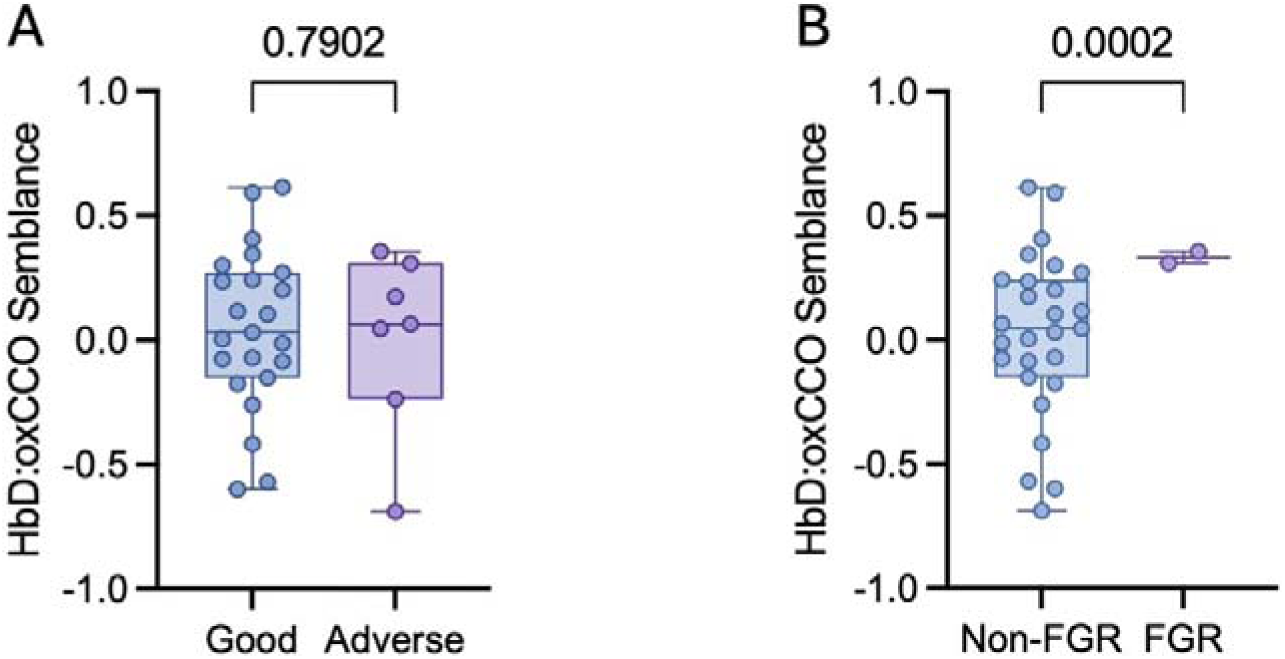
Mean HbD:oxCCO semblance between normal (n=22) and adverse (n=8) pregnancy outcome groups (p=0.7902) based on the near-miss criteria for stillbirth. (B). Mean PltO_2_ in pregnancies complicated by severe-FGR (n=3) and without (n=27) (C). Adverse = pregnancies that had a near-miss criteria for stillbirth.

**Figure 6.**
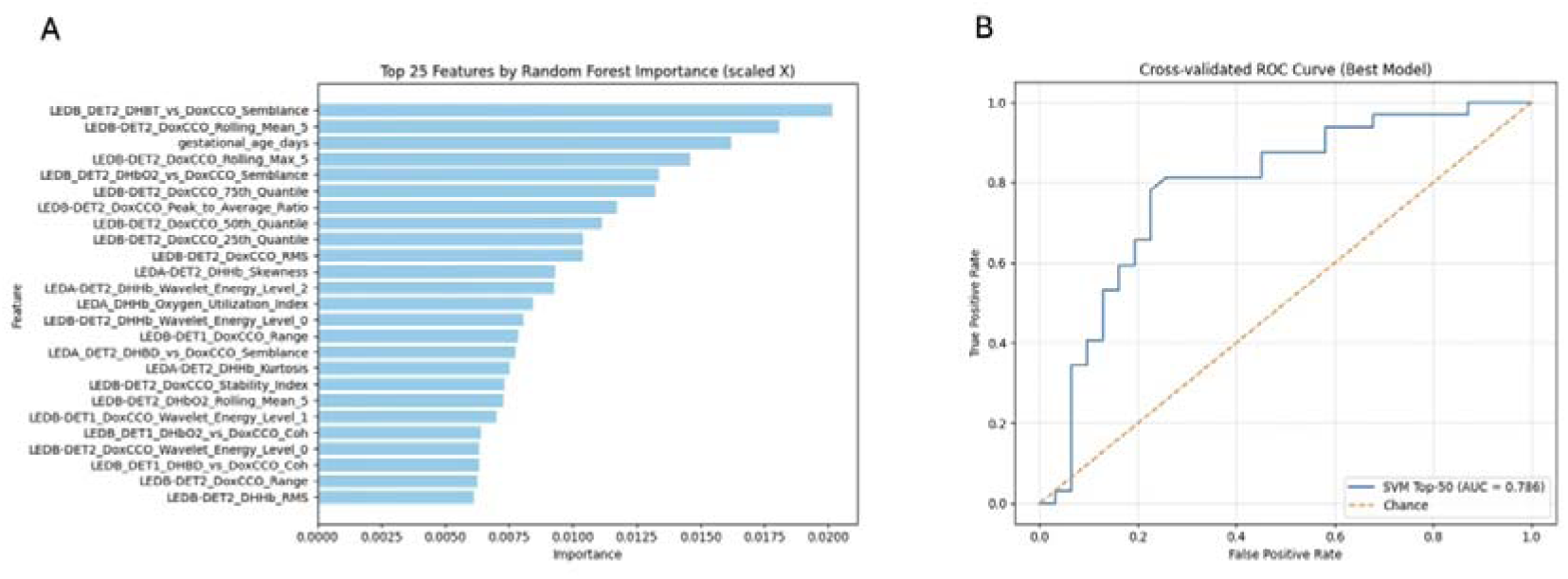
Top 25 features by random forest importance in the ML analysis, indicating a significant contribution from oxCCO related features (A). Receiver operating characteristic curve showing the AUC of 0.786 for the best-performing model (SVM with 50 features).

### Machine learning analysis

After preprocessing and DBSCAN-based outlier removal, 11 classifiers were tested, and the SVM model with the top 50 features performed best in pregnancy outcome prediction, achieving a balanced accuracy of 78%, recall (sensitivity) of 72% and specificity of 84% under 5 x 4 nested cross-validation.

Feature importance analysis revealed a strong contribution of metabolic features to outcome prediction. OxCCO-derived variables dominated the highest-ranking predictors, with haemodynamic measures playing a lesser role. In particular, semblance features capturing the temporal coupling between HbD and oxCCO (the relationship between oxygenation-related haemodynamic changes and metabolic function) consistently appeared among the top predictors. In contrast, static indices such as PltO_2_ had limited discriminative power, consistent with the lack of significant associations observed in univariate analysis. These results indicate that ML applied to placental NIRS signals can achieve high predictive accuracy, and that metabolic activity measured by oxCCO is a key driver of model performance.

In the MPS sub-cohort, the extra trees model with top 20 features demonstrated the best performance, achieving 80% for each of the key performance metrics: balanced accuracy, recall (sensitivity), and specificity under 5 x 4 nested cross-validation.

## Comment

### Principal findings

In this prospective observational study, we evaluated a wearable CW-NIRS system capable of simultaneously monitoring placental oxygenation and metabolism in vivo.

This study presents the first wearable CW-NIRS prototype capable of simultaneously monitoring placental oxygenation and metabolism. FSM v1 generated clinically usable data, including in participants with posterior placentas, confirming the feasibility of in vivo transabdominal monitoring across a heterogeneous cohort of high-risk pregnancy participants.

Static mean placental oxygenation values did not differ by pregnancy outcome; however, this might be related to small sample sizes. We found pregnancies complicated by FGR had higher PltO_2_ values, aligning with findings from previous NIRS studies^6,8,12^. In contrast to FGR pregnancies, those affected by GDM demonstrated lower PltO_2_ values. Interestingly, we also found higher HbD:oxCCO semblance values in severe FGR, indicating likely metabolic dysfunction, although larger sample sizes are required to validate these findings.

Our ML analysis revealed that classifiers trained on dynamic features (including frequency-domain and wavelet-derived) predicted adverse pregnancy outcomes for near miss stillbirth criteria with high accuracy. The best performing model, the support vector machine (SVM), achieved a balanced accuracy of 78%. Feature importance analysis provided mechanistic insight. Metabolic features dominated the predictive ability of the ML models, with oxCCO-derived variables consistently ranked amongst the most influential. Semblance features, quantifying the degree of coupling between HbD and oxCCO, also featured prominently.

### Results in the context of what is known

Earlier CW-NIRS prototypes were largely limited to anterior or fundal placentas and short measurement windows. For example, *Kakogawa et al*. demonstrated feasibility of placental issue oxygenation index (TOI) measurements using a commercial Hamamatsu NIRO device, but only in anterior placentas and small, condition-specific cohorts such as PET or FGR^6,7,23^. Similarly, *Razem et al.* reported intrapartum NIRS monitoring, but again restricted to anterior and fundal placentas, limiting wider applicability^24^. These studies illustrate both technical feasibility and the methodological constraints that FSM v1 was designed to overcome. Inconsistencies in the literature with regards to NIRS signatures in different placental pathologies likely reflect both methodological constraints and the limitations of static indices that fail to capture the complex temporal interplay between oxygen delivery and utilisation.

Across placental NIRS studies, reported findings have been inconsistent, with both reduced and paradoxically elevated placental oxygenation described in pregnancies complicated by FGR or placental insufficiency^6–8,12,25^. Placental NIRS measurements represent a composite signal that is likely dominated by maternal haemoglobin within the intervillous space (IVS), given its large blood volume, slower flow and proximity to the maternal abdominal wall ^12^. Under conditions of impaired maternal-fetal exchange such as FGR, reduced villous surface and oxygen extraction can result in relative oxygen accumulation within the IVS, yielding higher measured placental oxygenation despite evidence of fetal compromise and adverse outcomes, consistent with prior NIRS and MRI studies. Notably, these findings become more pronounced within the MPS sub-cohort, supporting the premise that adequate placental signal contribution might be useful for better physiological understanding. This reinforces the importance of incorporating optical modelling into placental NIRS studies, particularly when monitoring across heterogeneous maternal body habitus, placental positions, maternal skin tone and placental depths.

In contrast to FGR pregnancies, those affected by GDM demonstrated lower PltO_2_ values. While the sample sizes are small, this pattern may reflect altered placental metabolic and vascular physiology in the context of maternal hyperglycaemia. Human placental studies have demonstrated significantly increased release of oxidative stress markers and elevated activity of antioxidant enzymes in placentas from women with GDM compared with healthy controls^27^. These changes may reflect increased placental oxygen consumption and altered mitochondrial function in the hyperglycaemic environment. Whereas the observed elevated PltO_2_ in severe FGR likely indicates impaired oxygen extraction, underscoring that placental NIRS signals reflect the balance between oxygen delivery and utilisation, rather than simply perfusion alone. These findings should be interpreted with caution due to small sample sizes but are useful in demonstrating potential proof-of-concept.

The emergence of metabolism as a dominant predictor for adverse pregnancy outcomes aligns with evidence from both preclinical placental studies and neonatal cerebral monitoring, where oxCCO has been shown to reflect the adequacy of oxygen utilisation at the cellular level^4,5,28,29^. The relationship between oxygenation-related haemodynamic changes and mitochondrial utilisation (rather than absolute oxygenation) carries the greatest diagnostic weight. From a physiological perspective, this is compelling: mitochondrial dysfunction is the final common pathway in placental insufficiency, and disturbances in oxidative metabolism may precede or better reflect pathological states than haemodynamic measures alone. This has been demonstrated in multiple studies with decreased ATP production, increased reaction oxygen series (ROS) and abnormal mitochondrial dynamics observed in conditions associated with placenta insufficiency (such as PET and FGR)^30^.

### Clinical and research implications

These findings have important implications. Firstly, they provide a rationale for shifting research focus away from static haemoglobin indices towards dynamic metabolic monitoring, supported by advanced analytics. Second, they highlight the potential of ML-driven NIRS biomarkers as novel tools for advanced risk stratification. Third, they demonstrate that placental NIRS is not redundant simply because univariate studies have yielded inconclusive results so far; rather, its value lies in the complexity of the signal and the need for more appropriate analytical frameworks. Lastly, most previous studies have used commercial systems, which were not designed for use in the placenta or deep abdominal organs.

It is important to recognise that these insights are specific to NIRS-based monitoring. Unlike MRI, which can provide spatially resolved maps of placental oxygenation, NIRS has inherently limited spatial resolution and interrogates a heterogeneous placental tissue volume beneath the probe.

Integration of NIRS features with complementary modalities, including Doppler ultrasound, maternal serum biomarkers and digital CTG analytics, represents another promising avenue for multimodal risk prediction.

### Strengths and limitations

Our work has several limitations. Placental histopathology was only available in a subset of cases and thus was not included for final analysis. FSM v1 is portable; however, it requires a wired connection to a laptop and, therefore, cannot yet be considered a fully wearable system. Future iterations should focus on miniaturisation, wireless transmission and enhanced depth sensitivity (particularly for posterior placentas). Given the limited sample size, ML performance estimates should be interpreted as proof-of-principle rather than definitive predictive accuracy.

In addition, the 5% minimum placental sensitivity threshold was empirically derived from modelling outputs and should be refined and validated in larger, independent cohorts.

While our cohort was restricted to a single London centre, its socioeconomic and ethnic diversity provides a valuable foundation. Broad validation in multi-centre, multi-ethnic populations will be necessary to confirm generalisability and to develop algorithms suitable for clinical use. Lastly, our study represents a population within a high-income setting, and future studies should aim to study the utility of placental NIRS in low- and middle-income settings, where there is the greatest clinical need.

## Conclusion

FetalSenseM v1 represents the first wearable NIRS system capable of simultaneous monitoring of placental oxygenation and metabolism. In a high-risk cohort, we demonstrated feasibility across different placental positions, generating robust signals in most participants. Higher mean PltO_2_ values were observed in severe FGR and lower values in GDM, indicating the utility of FSM v1 in identifying different pathophysiological states. However, when predicting adverse pregnancy outcome, such as near miss for stillbirth, static indices, such as PltO_2_ were of limited diagnostic value, but when analysed with ML, dynamic haemodynamic and metabolic features (particularly oxCCO and its coupling to haemodynamic oxygen-related signals) predicted pregnancy outcome with high accuracy. These findings establish the principle that continuous placental NIRS, coupled with advanced analytics, can generate clinically meaningful biomarkers of placental function. Larger multi-centre studies are now warranted to validate predictive algorithms and refine device performance, with the ultimate goal of integrating wearable placental monitoring with advanced analytics into routine antenatal care.

## Funding

The authors were supported by the Wellcome Trust (219610/Z/19/Z), Wellcome Leap as part of the In Utero programme, Wellcome / EPSRC Centre for Interventional and Surgical Sciences (WEISS), and National Institute for Health Research University College London Hospitals Biomedical Research Centre.

## Author contributions

SM conceptualised the study. IT, TI, MT and UH conducted hardware and software developments. NRZ collected clinical data with support from SJ, ON, DS, SH, ALD and SM. NRZ and SM conducted the initial analysis, and statistical guidance was provided by HD. ZS conducted machine learning analysis with support from YK. SM and NRZ developed the first draft. All authors reviewed, contributed to and approved the final manuscript.

## Conflict of Interest

None

## Data Availability

All data produced in the present study are available upon reasonable request to the author

## Notes

### Competing Interest Statement

The authors have declared no competing interest.

### Author Declarations

Ethics Committee of East of England - Essex Research Ethics Committee gave ethical permission for this work.

