## Supplementary material for "Antenatal surveillance of placental function using a wearable near infrared spectroscopy device with machine learning data interpretation": Table 1

| Maternal Age (years) | 36.1 (±5.5) |
| --- | --- |
| BMI | 28.6 (±6.2) |
| Primiparous | 33 (56.9%) |
| Gestational age (weeks + days) at recruitment | 33+5 (± 3+6) |
| **Ethnicity (n (%))**  White (British, Irish, European, Other)  Other  Asian – Indian  Asian – Bangladeshi  Asian – Pakistani  Asian - Other  Chinese  Mixed - White and Asian  Mixed – White and Black African or Caribbean  Black – African, Caribbean or Other | 32 (55.2%)  4 (6.9%)  5 (8.6%)  3 (5.2%)  3 (5.2%)  1 (1.7%)  2 (3.4%)  1 (1.7%)  1 (1.7%)  3 (5.2%) |
| **Mode of birth**  Spontaneous vaginal birth  Instrumental birth(Forceps or Vacuum)  Emergency Caesarean section  Elective Caesarean section  Caesarean section (all) | 12 (20.7%)  3 (5.2%)  25 (43.1%)  18 (31.0%)  43 (74.1%) |
| **Clinical outcomes**  Good  Adverse: positive for near-miss criteria for stillbirth  **Reasons for adverse outcomes**  Severe PET  Severe FGR  High grade MVM  Neonatal CPR | 28 (48.2%)  30 (51.7%)  11 (18.9%)  13 (22.4%)  2 (3.4%)  1 (1.7%) |
| **Placental position**  Anterior  Posterior  Fundal | 29 (50.0%)  27 (46.5%)  2 (3.4%) |

**Table 1.** Study participant characteristics for the full participant cohort (n=58). PET: pre-eclampsia; FGR: fetal growth restriction; MVM: maternal villous malperfusion in the placenta; CPR: cardiopulmonary resuscitation.
